# Circular RNA expression profiles in pediatric ependymomas

**DOI:** 10.1101/2020.08.04.20167312

**Authors:** Ulvi Ahmadov, Meile M. Bendikas, Karoline K. Ebbesen, Astrid M. Sehested, Jørgen Kjems, Helle Broholm, Lasse S. Kristensen

**Affiliations:** Department of Biomedicine, Aarhus University, Aarhus, Denmark; Molecular Biology and Genetics (MBG), Aarhus University, Aarhus, Denmark; Interdisciplinary Nanoscience Center (iNANO), Aarhus University, Aarhus, Denmark; Department of Pediatrics and Adolescent Medicine, Copenhagen University Hospital, Copenhagen, Denmark; Department of Pathology, Center of Diagnostic Investigation, Rigshospitalet, Copenhagen, Denmark

**Keywords:** Pediatric ependymoma, pilocytic astrocytoma, medulloblastoma, circular RNA, RNA-sequencing, NanoString nCounter

## Abstract

Pediatric brain tumors frequently develop in the cerebellum, where ependymoma, medulloblastoma and pilocytic astrocytoma are the most prevalent subtypes. These tumors are currently treated using non-specific therapies, in part because few somatically mutated driver genes are present, and the underlying pathobiology is poorly described. Circular RNAs (circRNAs) have recently emerged as a large class of primarily non-coding RNAs with important roles in tumorigenesis, but so far they have not been described in pediatric brain tumors. To advance our understanding of these tumors, we performed high-throughput sequencing of ribosomal RNA-depleted total RNA from 10 primary ependymoma and 3 control samples. CircRNA expression patterns were determined using two independent bioinformatics algorithms, and correlated to disease stage, outcome, age, and gender. We found a profound global downregulation of circRNAs in ependymoma relative to control samples. Many differentially expressed circRNAs were discovered and circSMARCA5 and circ-FBXW7, which are described as tumor suppressors in glioma and glioblastomas in adults, were among the most downregulated. Moreover, patients with a dismal outcome clustered separately from patients with a good prognosis in unsupervised hierarchical cluster analyses. Next, we performed NanoString nCounter experiments using a custom-designed panel including 66 selected circRNA targets and analyzed formalin-fixed paraffin-embedded (FFPE) samples from a larger cohort of ependymoma patients as well as patients diagnosed with medulloblastoma or pilocytic astrocytoma. These experiments were used to validate our findings and, in addition, indicated that circRNA expression profiles are different among distinct pediatric brain tumor subtypes. In particular, circRMST and a circRNA derived from the *LRBA* gene were specifically upregulated in ependymomas. In conclusion, circRNAs have profoundly different expression profiles in ependymomas relative to controls and other pediatric brain tumor subtypes.

## Introduction

Ependymomas are rare cancers of the central nervous system (CNS), which mostly occur intracranially (supratentorial brain and posterior fossa) in children between 0 and 4 years of age, but they are also observed in older children and adults. Clinical management is challenging, and pediatric patients with intracranial ependymomas have high mortality rates. Surgical resection combined with radiotherapy remains the standard-of-care treatment and prediction of patient outcome based on tumor location, clinical characteristics and histopathology is challenging^1, 2^. This is mainly due to a significant variance in the grade II versus grade III distinction, even between experienced neuropathologists^3^, and tumors with histopathological similarities are heterogeneous at the molecular level resulting in diverse clinical outcomes.

In the majority of cases, the underlying oncogenic drivers are unknown and the underlying pathobiology of ependymoma is poorly described. This may, in part, be attributed to a very low mutation rate and problems in establishing ependymoma cell lines and animal models^4^. So far, only the C11orf95-RELA fusion, which results from chromothripsis and drives oncogenic NF-κB signaling^5^, has been incorporated into the World Health Organization (WHO) classification of CNS tumors^6^. This fusion gene characterizes more than 70% of the supratentorial ependymomas and is associated with poor outcome^2^. A better molecular understanding of the pathobiology of the disease may assist the development of targeted therapies and lead to the discovery of better prognostic markers.

Recently, circular RNAs (circRNAs) have emerged as a large class of endogenous RNAs with mainly non-coding functions^7, 8^, which play key roles in development and disease^9, 10^. They exhibit tissue-specific expression patterns and constitute a significant amount of cellular RNA, particularly in the brain^11-14^. Importantly, these molecules are extremely stable^7, 15, 16^ and situated in the cytoplasm where they may bind other cellular molecules, such as microRNAs (miRs)^7, 17^ or proteins^18-20^, and regulate their functions.

In pediatric brain tumors, including ependymoma, nothing is currently known about the expression and potential deregulation of circRNAs. On the other hand, circRNAs are emerging as important oncogenic drivers and tumor suppressors in glioma and glioblastoma, mainly by functioning as miR sponges^21-25^, protein sponges^26^ or as templates encoding tumor suppressor proteins^27-29^ through cap-independent translation via internal ribosome entry sites (IRESs).

In the present study, we employed high-throughput RNA-sequencing (RNA-seq) for an unbiased identification and profiling of circRNAs. Using this approach, we were able to describe the genome-wide circRNA expression landscape in pediatric ependymoma. We delineated expression differences between ependymomas and non-malignant control brain tissues as well as between long-term survivors and deceased patients. The use of RNA-seq also allowed us to investigate whether differentially expressed circRNAs were changed independent of their cognate linear host genes. To validate and further investigate circRNA expression profiles in ependymoma, we analyzed the expression of 66 selected circRNAs in a second independent cohort consisting of ependymomas, pilocytic astrocytomas, medulloblastomas and controls using the NanoString nCounter technology.

## Materials and Methods

### Patient and control samples

A cohort consisting of ten pediatric patients diagnosed with ependymoma and three control fresh frozen samples yielding high quality RNA were used for a genome-wide discovery study of circRNA expression profiles using RNA-seq. Clinical data were extracted from the patient files and critically reviewed. Patient characteristics are summarized in Table 1. An additional cohort consisting of 19 pediatric ependymoma, five pilocytic astrocytoma and three medulloblastoma patients as well as nine controls from formalin-fixed paraffin-embedded (FFPE) tissues were studied to validate circRNA expression changes found in the RNA-seq study. All patients were diagnosed at Rigshospitalet in Copenhagen, Denmark according to the applicable WHO guidelines and all samples were interrogated by an experienced pathologist to confirm that they contained at least 80% cancer cells.

**Table 1.** Clinical characteristics of the ependymoma patients used for RNA-seq.

| Patient number | Anatomical site | WHO grade | Age at diagnosis | Clinical follow-up time (years) | Survival status |
| --- | --- | --- | --- | --- | --- |
| 1 | Fossa posterior | II | 2 | 3 | Deceased |
| 2 | Fossa posterior | II | 5 | 20 | Alive |
| 3 | Fossa posterior | II | 5 | 20 | Alive |
| 4 | Fossa posterior | II | 2 | 9 | Unknown |
| 5 | Fossa posterior | II | 1 | 3 | Deceased |
| 6 | Fossa posterior | II | 13 | 17 | Alive |
| 7 | Fossa posterior | III | 8 | 22 | Alive |
| 8 | Fossa posterior | III | 2 | 3 | Deceased |
| 9 | Fossa posterior | III | 2 | 6 | Deceased |
| 10 | Fossa posterior | III | 1 | 17 | Deceased |

### Ethical approval

This study has been conducted according to the Declaration of Helsinki and Danish legislation and approval from the national ethics committee has been granted (approval number: 1707758).

### RNA isolation

RNA from fresh frozen samples was isolated using the RNeasy Mini Kit (250) (Qiagen, Hilden, Germany), and RNA from formalin-fixed paraffin-embedded (FFPE) tissues was isolated using the Maxwell® RSC RNA FFPE Kit (Promega Corporation, Madison, WI, USA) according to the manufacturers’ instructions. The quantity and purity of total RNA was measured using a NanoDrop-1000 spectrophotometer (Thermo Scientific, Delaware, USA).

### RNA-seq library preparation, Illumina sequencing and initial data processing

One microgram of total RNA was rRNA depleted using the Ribo-Zero rRNA Removal Kit (Human, Mouse, Rat) (Epicentre, Madison, WI, USA) followed by a purification step using AMPure XP Beads (Beckman Coulter, Brea, CA, USA). Sequencing libraries were generated using the ScriptSeq v2 RNA-Seq Library Preparation Kit (Epicentre) using 12 PCR cycles for amplification. Purification was performed using AMPure XP Beads (Beckman Coulter). The final libraries were quality controlled on a 2100 Bioanalyzer (Agilent Technologies, Santa Clara, CA, USA) and quantified using the KAPA library quantification kit (Kapa Biosystems, Wilmington, MA, USA). RNA-seq was performed on the HiSeq 4000 system (Illumina, San Diego, CA, USA) at the Beijing Genomics Institute (BGI) in Copenhagen using the 100 paired-end sequencing protocol with twelve samples pooled on one lane. The resulting data were demultiplexed, quality filtered (Phred score 20) and adapter trimmed using Trim Galore.

### circRNA quantification in the RNA-seq

The filtered and trimmed sequencing reads were mapped to the human genome (hg19) using Bowtie2, mapping only unspliced reads. The unmapped reads were analyzed using a stringent version of the find_circ bioinformatics algorithm^12^ and the CIRCexplorer algorithm^30^. All analyses were based on the stringent find_circ algorithm, but circRNA candidates not detected by CIRCexplorer were manually inspected to exclude obvious mapping artifacts as previously described^31^. Reads per million (RPM) refers to sequencing reads aligning across the particular backsplicing junction divided by the total number of reads in the particular sample multiplied by one million. The circular-to-linear (CTL) ratios were defined as the number of reads spanning the particular backsplicing junctions divided by the average linear reads spanning the splice donor- and splice acceptor sites of the same backsplicing junction plus one pseudo count (to avoid division by zero).

### mRNA quantification in the RNA-seq data

Sequencing reads were quality-filtered, and adaptor-trimmed as described above. Filtered and trimmed sequencing reads were mapped to hg19 using Tophat2 and featureCounts^32^ was used to quantify the number of reads mapping to annotated genes from Ensembl gene definitions release 71. Differential expression analysis was performed using the DESeq2 R package.

### NanoString nCounter custom CodeSet for circRNA analysis

A custom CodeSet of capture- and reporter probes was designed to target regions of 100 bp overlaying the BSJs of 66 circRNAs selected based on the RNA-seq data (Supplementary Table 1). In addition, ten linear reference genes (*GAPDH, ACTB, SF3B1, B2M, RPL19, PUM1, GUSB, IPO8, TBP* and *HPRT1*) were included in the CodeSet. Approximately 50 ng of total RNA from each sample was hybridized to the capture- and reporter probes for 20 hours and then analyzed on the nCounter(tm) SPRINT platform (NanoString Technologies, Seattle, WA, USA) according to the manufacturer’s instructions.

### NanoString nCounter data analyses

The raw data were processed using the nSOLVER 4.0 software (NanoString Technologies). Following import of the raw data, a positive control normalization was performed using the geometric mean of all positive controls with the exception of the control named F, as recommended by the manufacturer. Then, a second normalization was performed using the geometric mean of the eight linear reference genes (*GAPDH, ACTB, SF3B1, RPL19, PUM1, GUSB, IPO8* and *TBP*) having the lowest coefficient of variance percentage (%CV), before exporting the data to Excel (Microsoft Corporation, Redmond, WA, USA).

### Heatmaps, unsupervised hierarchical cluster analyses and principal component (PCA) analysis

Heatmaps, unsupervised hierarchical cluster analyses and PCA analysis were performed using R software version 4.0.0 with the following packages from Bioconductor (https://bioconductor.org/biocLite.R) installed: ComplexHeatmap, circlize, dendextend and RColorBrewer. The hierarchical cluster analysis using RNA-seq data was performed following a z-score transformation of the RPM values for each circRNA with the Euclidean distance calculation method. Heatmap for NanoString nCounter data was plotted following a z-score transformation of the normalized counts for each circRNA without clustering. The PCA analysis was performed using R software version 4.0.0 using the ggplot2 package. Before plotting the data, a log2 transformation of the normalized counts for each circRNA was performed.

### Statistical analyses

All statistical tests were performed using Prism 8 (GraphPad, La Jolla, CA, USA). The volcano plots were generated by one unpaired t test per gene individually without assuming consistent standard deviation and without correction for multiple testing. However, p-values, for which correction for multiple testing was performed using the Holm-Sidak method, were also calculated. Comparison between the gene expression levels of the groups was done using a Mann Whitney test, as the data were not normally distributed according to the D’Agostino & Pearson normality test. Simple linear regression was used to assess potential correlations between average CTL ratios and average RPM values, between gene expression (RPM) and circRNA expression (RPM), between fold changes in RPM and fold changes in CTL ratios employing an F test to investigate if the slope was significantly non-zero.

## Results

### Identification and characterization of circular RNAs in ependymoma and control samples

To reveal the circRNA expression landscape in ependymoma, we performed RNA-seq on fresh frozen samples of 10 intracranial ependymomas and 3 controls (Table 1). Using a stringent version of find_circ^12^, we detected 11,217 unique circRNAs supported by at least two BSJ-spanning reads in a single sample; 6,975 in the ependymomas and 8,479 in the controls (Fig. 1a and 1b) (Supplementary Table 2). Interestingly, the circRNAs were much more abundant in the control samples relative to the ependymomas (Supplementary Fig. 1).

**Figure 1.**
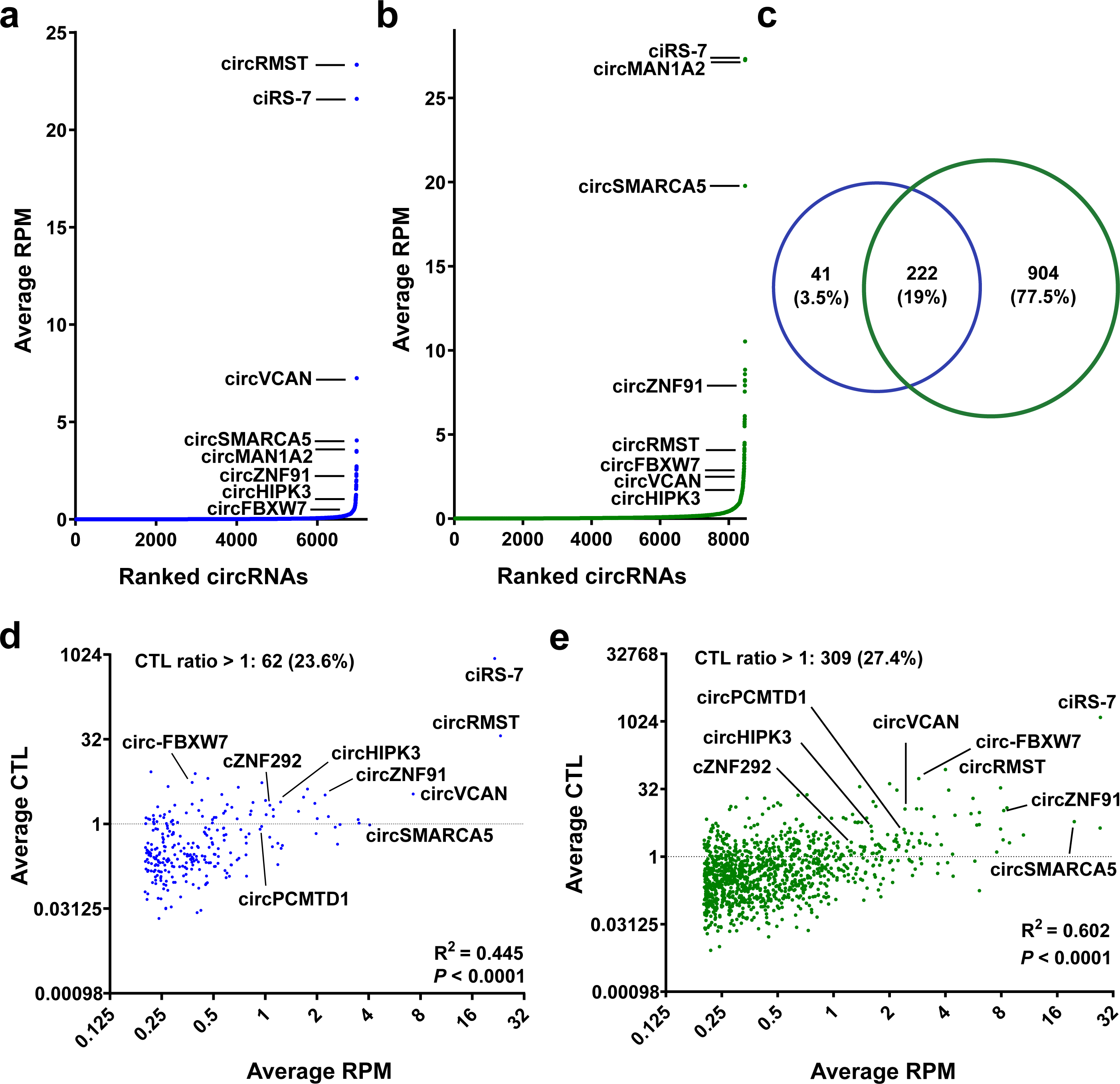
Identification of circular RNAs in ependymoma and control samples. **(a, b)** Scatter plots of unique circRNAs that are supported by at least two sequencing reads (BSJ >2) and ranked according to average expression level (average RPM) in ependymomas (**a**) and in healthy controls (**b**). (**c**) Venn diagram of high abundance circRNAs (average RPM >0.2) in ependymoma (blue circle) and healthy controls (green circle). **(d, e)** Correlation between average CTL ratios and average RPM values with corresponding linear regression statistics and R-squared values for the high abundance circRNAs in ependymomas (**d**) and in healthy controls (**e**). CircRNAs with a CTL ratio >1 are expressed at higher levels than their cognate linear host genes. Simple linear regression test is used to determine R and P-values. BSJ: backsplicing junction, CTL: Circular-to-linear, RPM: Reads-per-million.

CircRNAs supported by very few sequencing reads may not be biologically relevant and could potentially be reverse transcriptase- or sequencing artifacts^33, 34^. Therefore, we decided to analyze further only the most abundant circRNAs in the dataset. First, circRNAs with an average of at least 0.2 RPM values among either the ependymoma samples or the control samples were identified; 262 in the ependymomas and 1,126 in the controls. It should be noted on that the low number of circRNAs detected in ependymomas was not due to a difference in sample quality/sequencing depth. Most of these circRNAs, 231 of 263 (87.8%) in the ependymomas and 1033 of 1,126 (90.9%) (Supplementary Table 3), were also detected by CIRCexplorer, which detects circRNAs based on annotated splice sites. There was a substantial overlap (222 circRNAs) between the circRNAs detected in the ependymomas and in the controls and 41 and 904 were unique to the ependymomas and to the controls, respectively (Fig. 1c). Among the high abundance circRNAs, 62 (23.6%) and 309 (27.4%) were on average expressed at higher levels than their cognate linear host genes (CTL ratio >1) in the ependymomas and in the controls, respectively (Fig. 1d and 1e). Among the high abundance circRNAs, two potentially novel ones were detected (not present in circBase^35^ and CIRCpedia v2^36^), namely a circRNA derived from the *SH3KBP1* gene on chromosome X (circSH3KBP1) and a circRNA antisense to the *LRRC55* gene on chromosome 11 (circLRRC55-as).

Among the high abundance circRNAs, several have previously been shown to be aberrantly expressed in adult brain tumors, including circHIPK3^21, 22^, ciRS-7 (CDR1as)^23, 24^, circPCMTD1^25^, circSMARCA5^26^, circ-FBXW7^29^, cZNF292^37^ and circVCAN^38^ (Fig. 1d and 1e). On the other hand, we did not detect the circRNA derived from LINC-PINT^27^, which has been proposed to encode a tumor suppressor protein in glioblastoma.

### Unsupervised hierarchical cluster analysis of circRNA expression profiles reveals two distinct ependymoma subgroups

We performed unsupervised hierarchical cluster analysis using all the 1,167 high abundance circRNAs detected in the ependymomas and in the controls and observed that the ependymomas clustered separately due to a marked downregulation of the majority of the circRNAs (Fig. 2). Interestingly, the circRNA expression profiles from deceased patients were distinct from the profiles of patients who survived. Thus, two sub-clusters were observed among the ependymoma samples.

**Figure 2.**
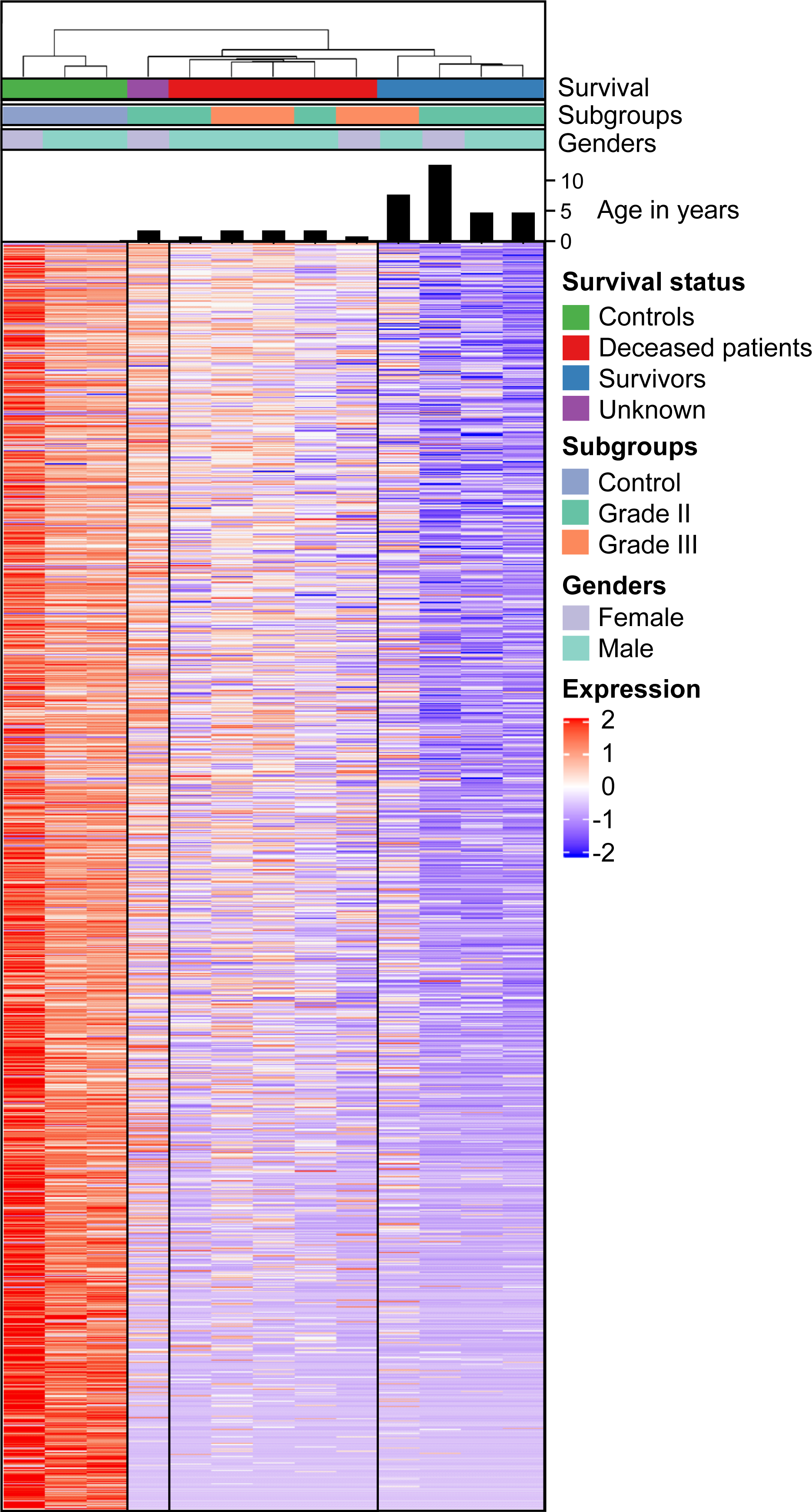
Unsupervised hierarchical cluster analysis of circRNA expression profiles reveals two distinct ependymoma subgroups. The heatmap and unsupervised hierarchical cluster analysis were created using the high abundance circRNAs (average RPM > 0.2) in ependymoma (n=10) and control (n=3) samples. This analysis revealed two distinct ependymoma subgroups. Age, WHO Grade (Grade II vs Grade III), survival status (survivors, deceased or unknown) and gender (male vs female) of the ependymoma samples are indicated. Each row in the heatmap corresponds to a unique circRNA quantified using RNA-sequencing. Each column corresponds to a patient or control sample. Each sample is annotated below the dendrogram in the top.

This sub-clustering according to outcome could not be explained by gender, tumor location (all the tumors were located in Posterior Fossa) or disease grade, but the deceased patients were diagnosed at a marked younger age (Fig 2). Pediatric Posterior Fossa ependymomas form two molecular subgroups based on DNA methylation analyses; one of these (PF-EPN-A) is characterized by a dismal outcome and a markedly younger median age^39, 40^. Therefore, it is highly likely that the subgroup with dismal outcome, which we identified by circRNA profiling, corresponds to PF-EPN-A. However, we were not able to confirm this as several attempts to perform 850K DNA methylation analyses failed due to poor DNA quality.

### Known trans-acting factors are unlikely to explain circRNA expression differences between the two ependymoma subgroups

Next, we analyzed the expression of genes that have previously been shown to directly affect backsplicing, in a search for potential trans-acting factors that may drive the observed clustering of the ependymoma samples into two groups. We focused on the protein quaking (protein product of *QKI*)^41^, RNA-binding protein FUS (protein product of *FUS*)^42^ and NF90/110 (protein products of *ILF3*)^43^, which have been shown to promote backsplicing, and ADAR1 (protein product of *ADAR*)^44^ and ATP-dependent RNA helicase A (protein product of *DHX9*)^45^, which inhibit backsplicing. Only *FUS* and *ADAR* expression levels were significantly different between the two groups (Supplementary Fig. 2a). When performing linear regression analyses, also including data from the controls, to assess possible correlations between the expression of each individual gene and overall circRNA expression levels in the samples, only *FUS* displayed a significant, albeit negative, correlation, while a non-significant positive correlation between *ADAR* and overall circRNA expression levels was observed (Supplementary Fig. 2b). Therefore, as *FUS* generally promotes backsplicing and *ADAR* inhibits backsplicing, none of the investigated trans-acting factors are likely to explain the overall reduction in circRNA abundance observed in the subgroup comprising all the survivors (Supplementary Fig. 2b and Supplementary Table 4).

### Genes involved in pre-mRNA splicing are unlikely to explain circRNA expression differences between the two ependymoma subgroups

Since it has been observed that protein-coding genes produce more circRNAs when the pre-mRNA processing machinery is limiting^46^, we decided to investigate a set of 280 genes involved in pre-mRNA splicing^47^ (Supplementary Table 5). Overall, there was no difference in the expression levels of these genes between the two subgroups nor between the survivors and the controls (Supplementary Fig. 3). When analyzing the genes individually, some were significantly downregulated and some significantly upregulated (Supplementary Fig. 4a and Supplementary Table 6). Moreover, in linear regression analyses, also including data from the controls, only five of these differentially expressed genes displayed a significant, albeit positive, correlation with overall circRNA expression levels (Supplementary Fig. 4b). Therefore, as protein-coding genes produce more circRNAs when the pre-mRNA processing machinery is limited, genes involved in pre-mRNA splicing events are unlikely to explain the overall reduction in circRNA abundance observed in the subgroup comprising all survivors.

### Key epigenetic modifier genes are differentially expressed between the two ependymoma subgroups

Next, we analyzed a set of 167 epigenetic modifier genes^48^ (Supplementary Table 7), since we and others have previously observed that epigenetics can influence circRNA expression patterns^15, 31, 49^. Among these genes, six were differentially expressed between the two subgroups; all being significantly more abundant in the subgroup comprising the survivors (Fig 3a and Supplementary Table 8). Moreover, three of these genes, *SETD4, INO80C* and *DNMT3B* showed a significant correlation in linear regression analyses, which also included data from the controls (Fig. 3b). Whether these genes have a direct effect on circRNA expression patterns in ependymoma cannot be inferred from these correlative studies, but we found it to be of particular interest that *DNMT3B* expression is correlated with overall circRNA abundance, since we have previously shown that *DNMT3B* knockdown results in circRNA expression changes that are independent of changes in their cognate linear host genes^31^. However, these previous experiments were done using epidermal stem cells and both upregulation and downregulation of circRNAs were observed upon knockdown of *DNMT3B*.

**Figure 3.**
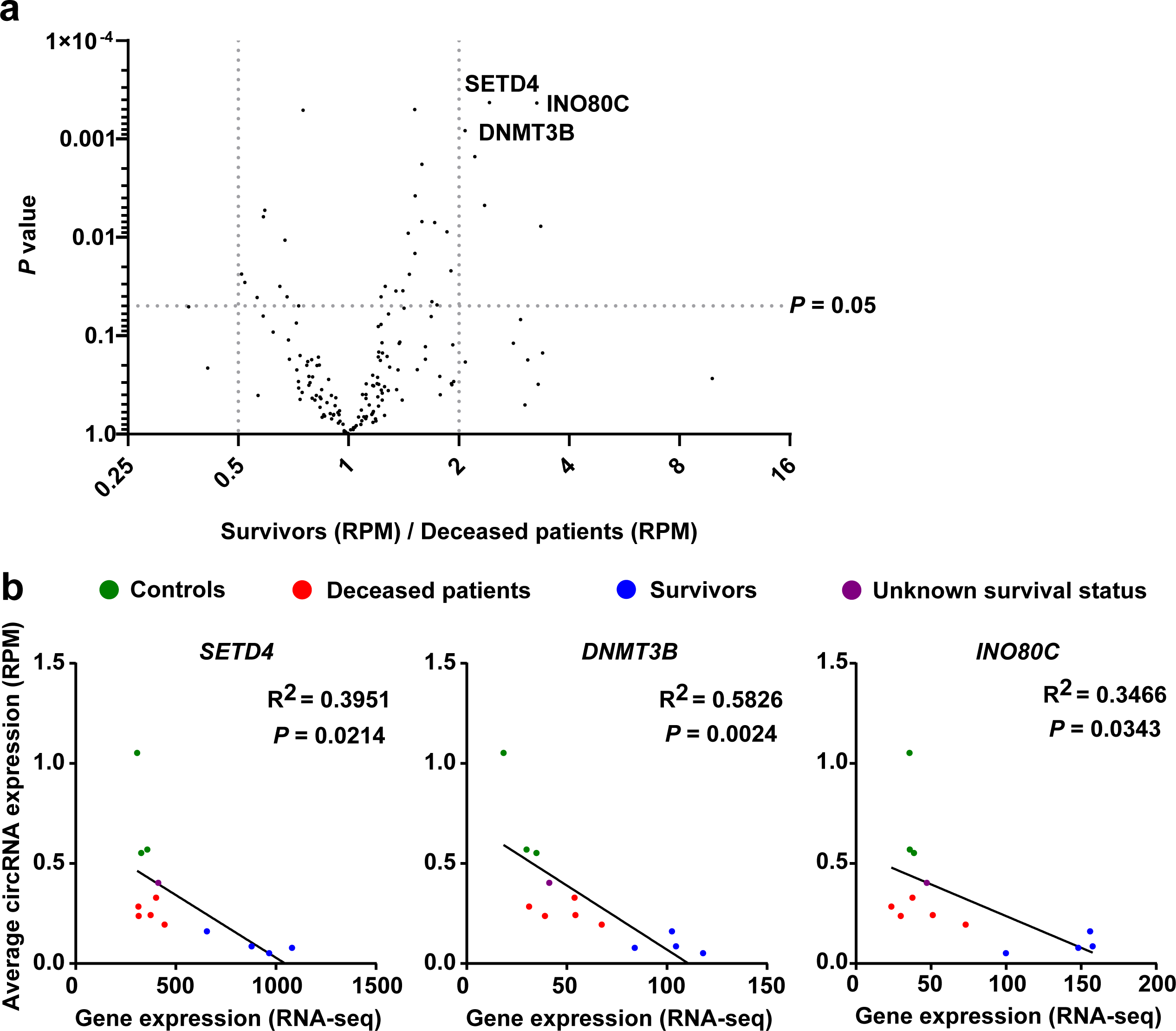
Key epigenetic modifier genes are differentially expressed between the two ependymoma subgroups. **(a)** Volcano plot of 167 epigenetic modifier genes showing several differentially expressed genes between survivors and deceased patients. One unpaired t test for each gene was used to generate the p-values. **(b)** Correlation between gene expression and circRNA expression (RPM) with corresponding linear regression statistics and R-squared values for the six significantly upregulated genes shown in panel a. Blue dots: samples from survivors: red dots: samples from deceased patients; green dots: samples from healthy controls; purple dot: sample from a patient with an unknown survival status. RPM: Reads-per-million.

### Differentially expressed circRNAs in ependymoma

Many of the circRNAs that displayed a marked downregulation in the ependymoma samples relative to the controls were statistically significant (Fig. 4a and Supplementary Table 9). The top 10 upregulated and downregulated circRNAs in ependymoma samples are listed in Table 2 and Table 3. Among the circRNAs which previously were shown to be aberrantly expressed in adult brain tumors, circSMARCA5 and circ-FBXW7 were found to be significantly downregulated in the ependymoma samples. This is consistent with the previous studies describing these circRNAs as tumor suppressors^26, 29^. Likewise, circVCAN and circRMST were more abundant, albeit not statistically significant, in ependymomas, similar to what has been observed in glioblastoma^38^. On the other hand, ciRS-7, circHIPK3, cZNF292 and circPCMTD1 were not aberrantly expressed in ependymoma (Supplementary Table 9). The circRNAs derived from the *ATRNL1* gene, which is highly expressed in brain tissues^50^, were some of the most downregulated circRNAs in ependymoma. In addition to downregulated circRNAs, the circRNAs derived from *DRC1, WDR49* and *VWA3A* genes were found to be significantly upregulated (Supplementary Table 9).

**Table 2.**
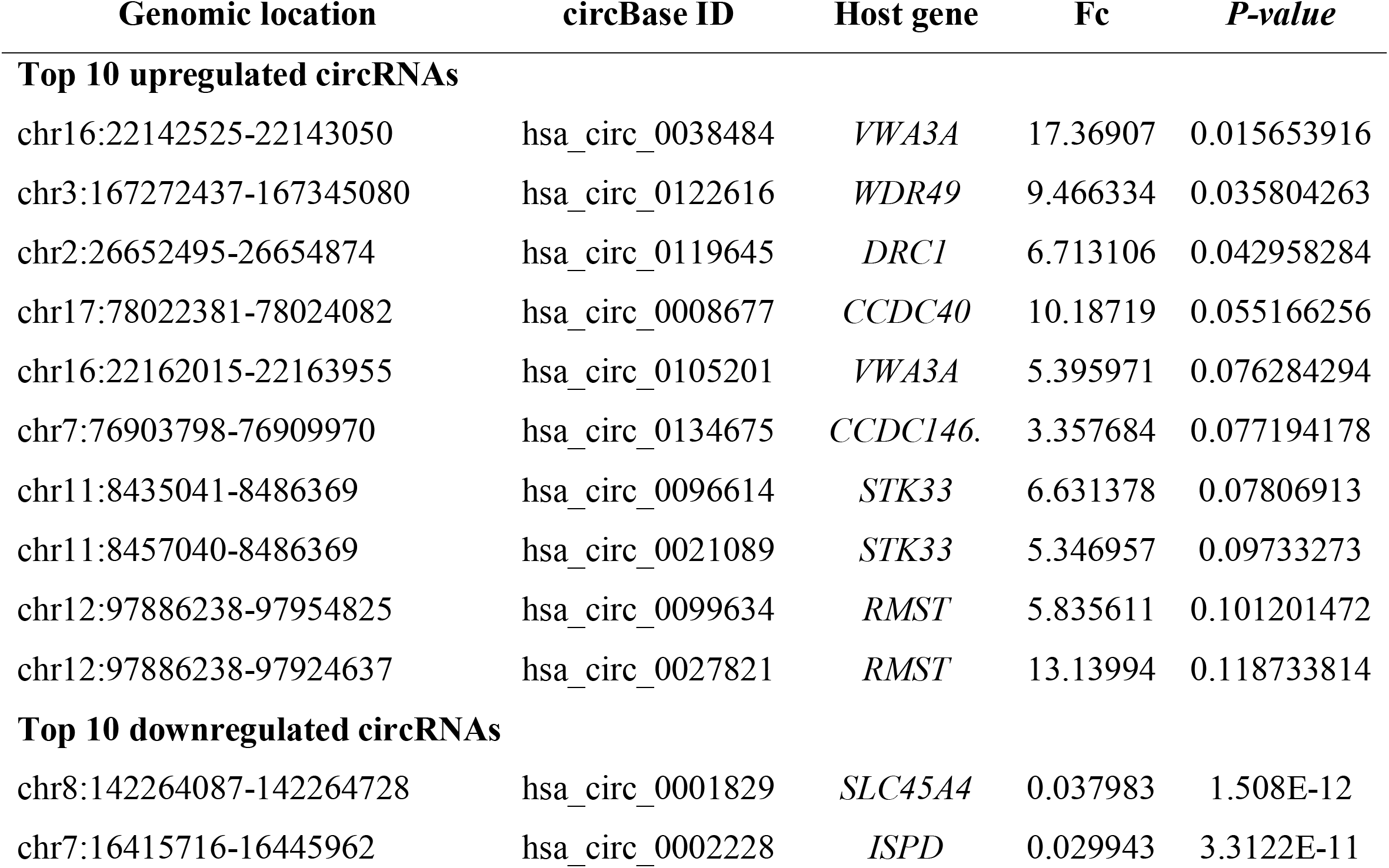
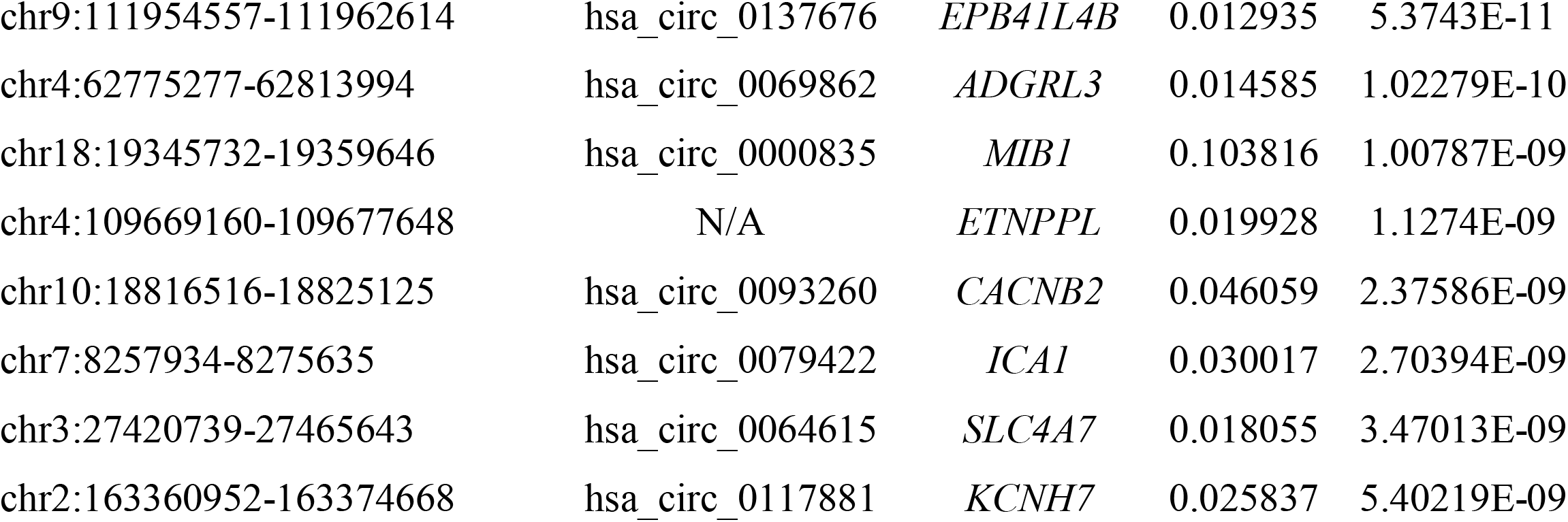
The top 10 most upregulated and downregulated circRNAs, respectively, in ependymoma samples relative to control samples. The circRNAs are listed according to their *P-value*s. Fc: fold change.

**Table 3.**
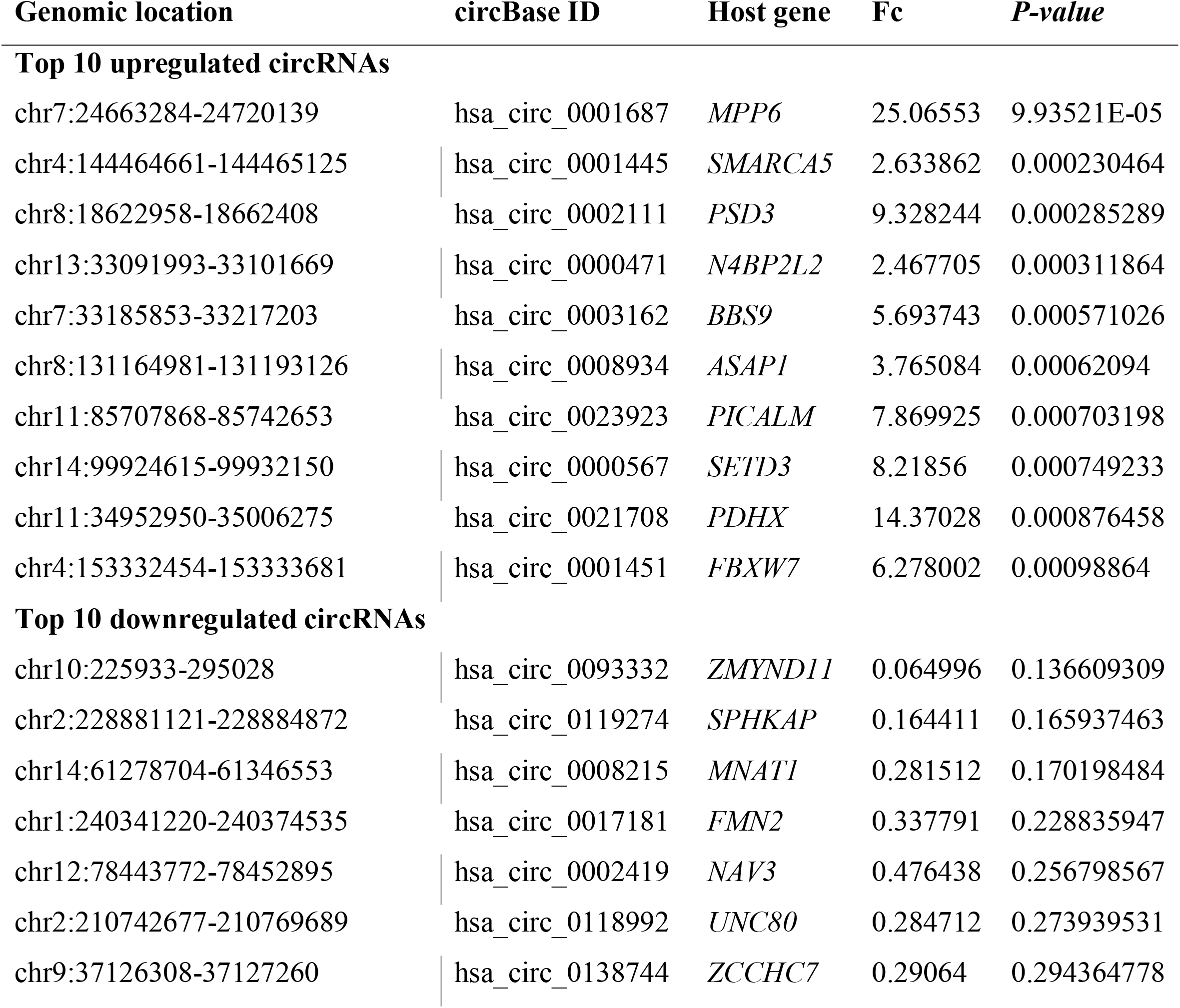
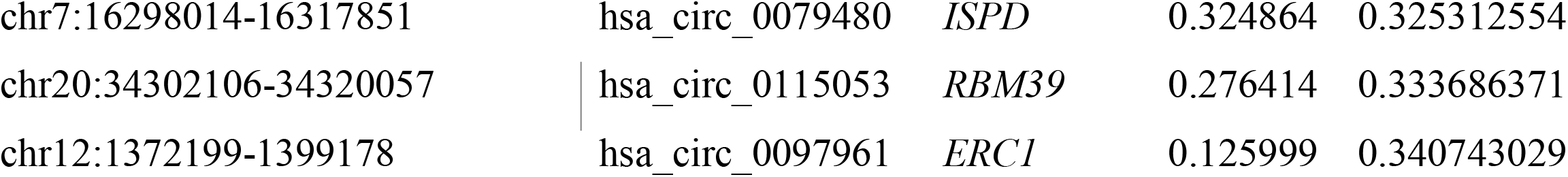
The top 10 most upregulated and downregulated circRNAs, respectively, in deceased ependymoma patients relative to ependymoma patients that survived. The circRNAs are listed according to *P-value*s. Fc: fold change.

**Figure 4.**
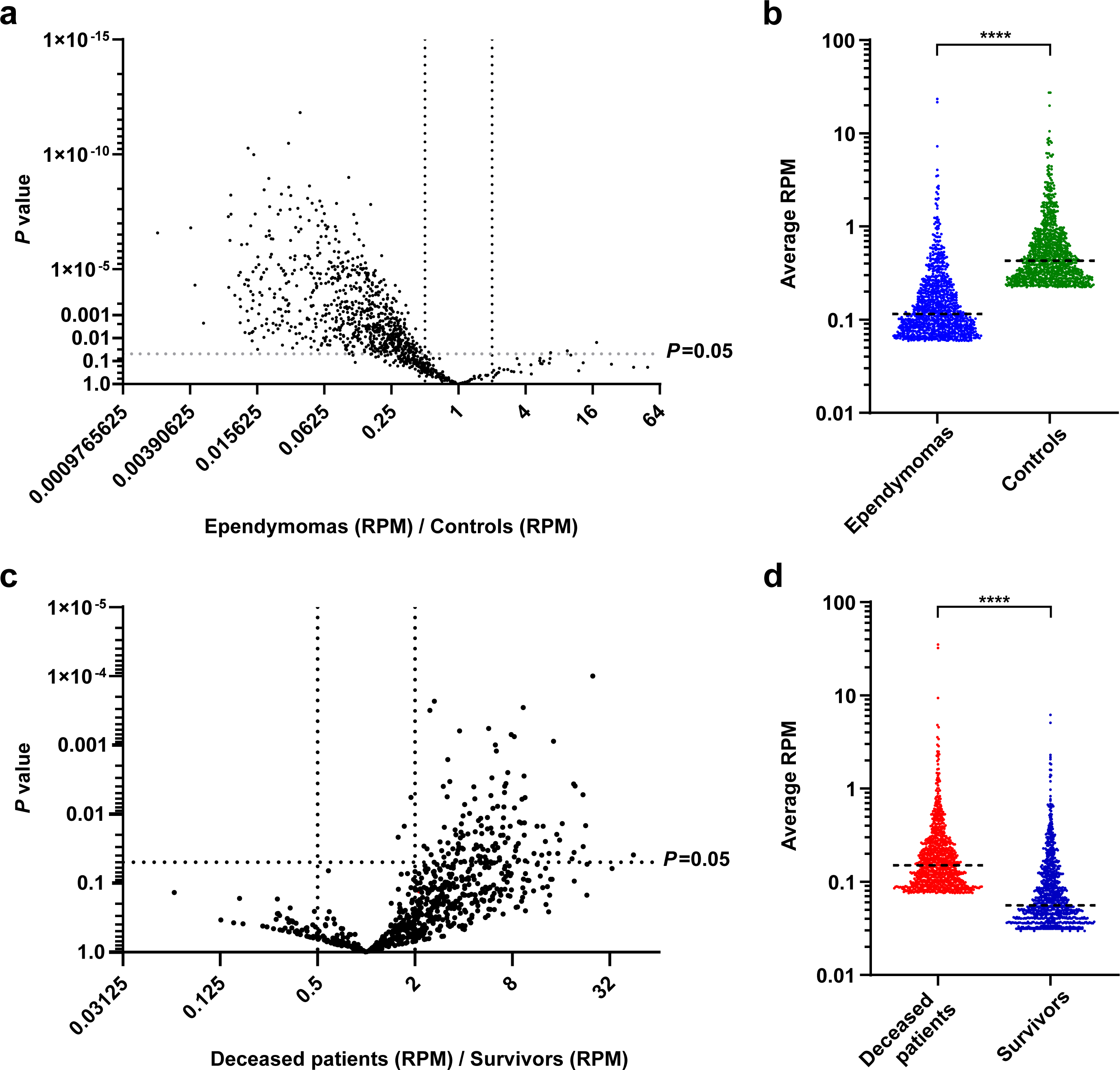
Expression profiling of differentially expressed circRNAs in ependymoma. **(a)** Volcano plot of 1167 high abundance circRNAs showing a high number of differentially expressed circRNAs between ependymoma (n=10) and control (n=3) samples. **(b)** Scatter plot of the top 1000 circRNAs identified in ependymoma (n=10) and control (n=3) samples. Mann Whitney U test is used for statistical calculation. **(c)** Volcano plot of 1167 high abundance circRNAs showing differentially expressed circRNAs between deceased patients (n=5) and survivors (n=4). **(d)** Scatter plot of the top 1000 circRNAs identified in samples from deceased patients (n=5) and survivors (n=4). Mann Whitney U test is used for statistical calculation. One unpaired t test for each circRNA was used to generate the p-values. * * * * *P-value* >0.0001

As expected from the heatmap (Fig. 2), we observed a significant overall upregulation of circRNAs for the deceased patients relative to the survivors when analyzing the data by a volcano plot (Fig. 4c and Supplementary Table 10) and a scatter plot (Fig. 4d). The top 10 upregulated and downregulated circRNAs is listed in Table 2 and Table 3. The upregulated circRNAs included cZNF292, consistent with a previous study describing an oncogenic role of cZNF292 in glioma^37^, circRMST and circ-FBXW7.

### Most differentially expressed circRNAs changed independent of their cognate linear host genes

To explore the relation between circRNA expression changes and potential expression changes of their cognate linear host genes, we plotted fold change (Fc) in circular-to-linear (CTL) ratios against Fc in reads-per-million (RPM) values. The majority of the differentially expressed circRNAs in the tumors relative to the controls, including cZNF292, circHIPK3, circPCMTD1, circSMARCA5 and circ-FBXW7, changed independent of their respective host genes (Supplementary Table 9 and 10) (defined as Fc (RPM)/ Fc (CTL) < 2; data points found between the dotted blue lines in figure 5a). On the other hand, the circRNAs derived from the *ATRNL1* gene were downregulated together with the linear cognates (Fig. 5a); consistent with a previous study showing that promoter CpG island hypermethylation of *ATRNL1* causes downregulation of both mRNA and circRNA^49^. Moreover, circVCAN, circRMST, and the circRNAs derived from *DRC1, WDR49* and *VWA3A* genes changed together with their linear cognates.

**Figure 5.**
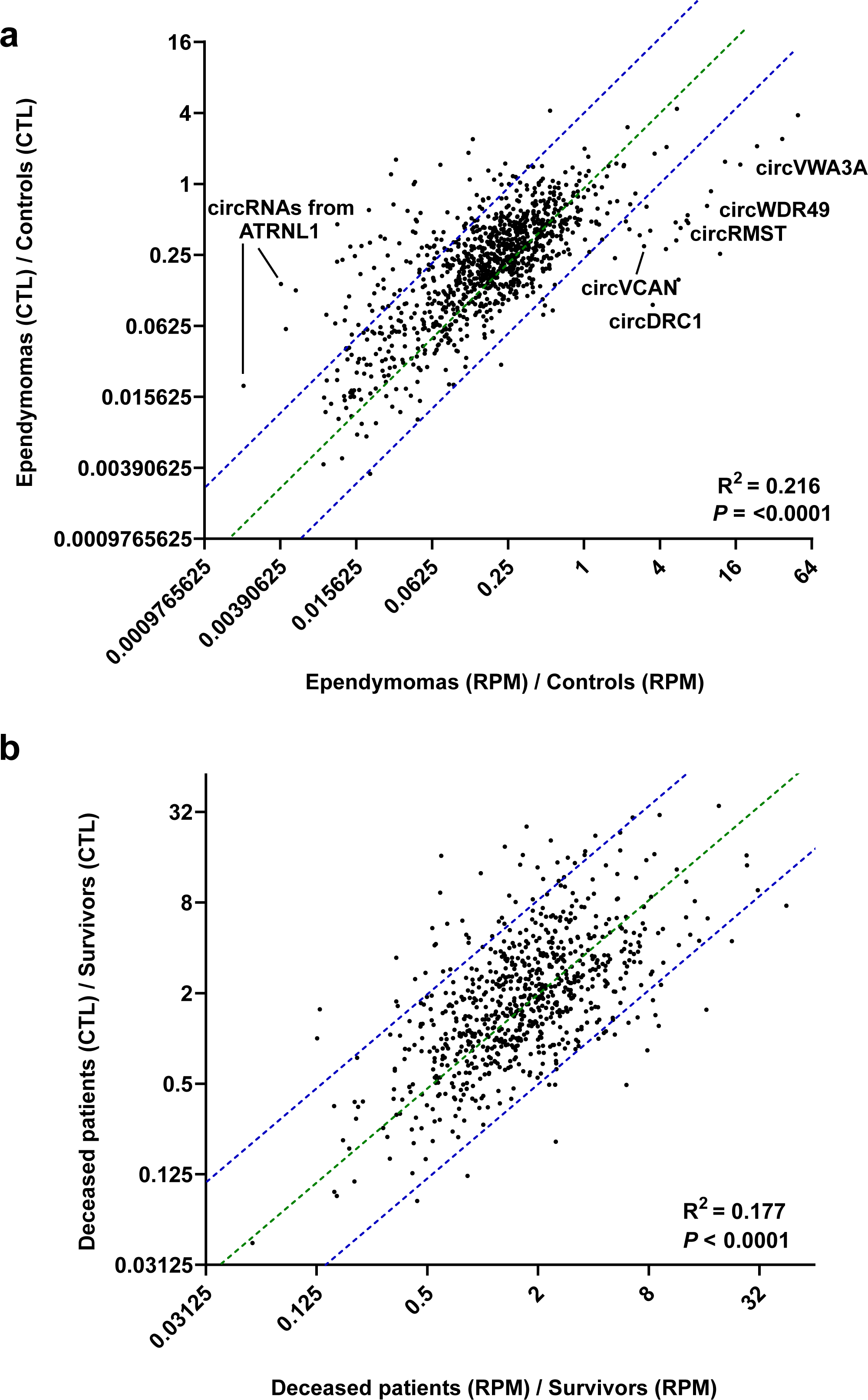
Most differentially expressed circRNAs were changed independent of their cognate linear host genes. **(a, b)** Scatter plots of fold change in CTL ratios against fold change in RPM values for circRNAs with corresponding linear regression statistics and R-squared values between ependymoma (n=10) and control (n=3) samples (**a**) and between deceased patients and survivors (**b**). CircRNAs between the dotted blue lines are considered changed independent of their respective host genes. CTL: Circular-to-linear, RPM: Reads-per-million.

When looking at differentially expressed circRNAs in the deceased patients relative to the survivors, a smaller fraction of the circRNAs were changed independent of their cognate linear host genes (Fig. 5b). In addition to circRNAs derived from *DRC1, VWA3A* and *WDR49* genes, circPCMTD1, circHIPK3, circVCAN, ZNF292, circRMST and circSMARCA5 changed independent of their respective host genes. Together, these analyses indicate that the majority of the differentially expressed circRNAs identified in figure 4a and 4c cannot be explained as passenger events due to gene expression changes in the cognate linear hosts.

### Validation of ependymoma-related circRNA expression changes using NanoString nCounter

To validate and further investigate circRNA expression profiles in ependymoma, we collected another independent cohort of 19 ependymomas as well as 5 pilocytic astrocytomas, 3 medulloblastomas and 9 control samples. Because these samples were derived from FFPE tissues, we decided to use the NanoString nCounter technology for circRNA quantification as this method has proved to work well for circRNA quantification in highly degraded RNA samples^51^. Moreover, this technology is not subject to bias and errors introduced by the use of enzymes, which is a particular concern in the circRNA research field^9, 33, 34, 52, 53^. Thus, we designed a custom NanoString nCounter probeset targeting 66 unique circRNAs (selected based on differential expression and abundance in the RNA-seq data) (Supplementary Table 1) and 10 potential reference genes for normalization of the data. First, as we expected based on how the circRNAs were selected, the ependymoma samples were distinguished from the control samples (Fig. 6) and the circRNAs were on average more abundant in control samples compared to ependymoma samples (Fig. 6 and Supplementary Fig. 5). In these analyses, we also observed that individual circRNAs derived from the same host genes tended to cluster together (e.g. circRNAs from *PSD3, SATB2* and *SLC8A1*) (Fig. 6). In addition, we observed a strong correlation between the NanoString nCounter data and the RNA-seq data; the correlation coefficient (R^2^) was 0.74 with p<0.0001 when comparing log_2_ Fc values between ependymoma and control samples (Supplementary Fig. 6). Since we analyzed two independent cohorts, one consisting of FFPE tissue samples and the other of fresh frozen samples, using two different methodologies, some variation is to be expected.

**Figure 6.**
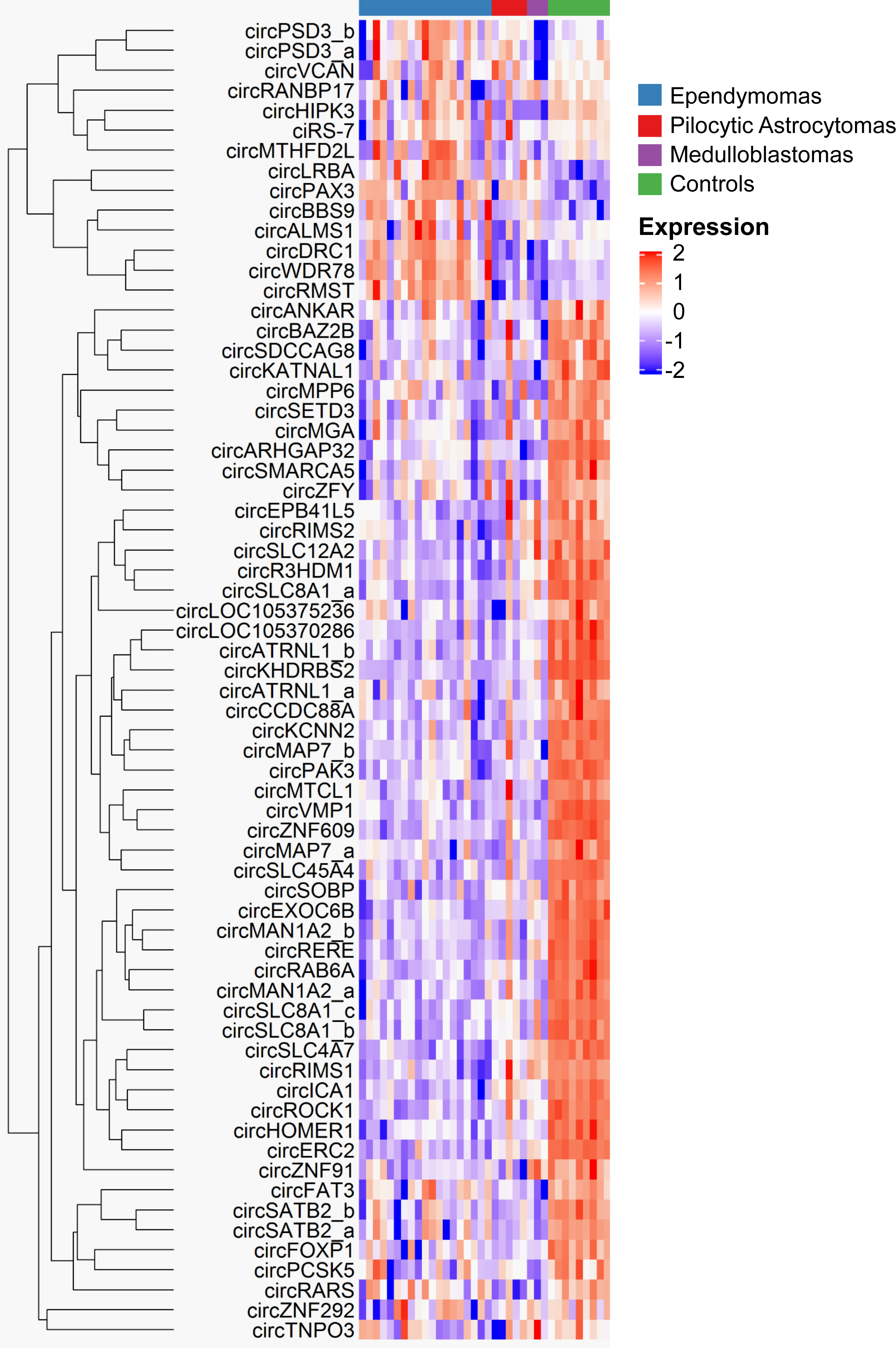
Validation of ependymoma-related circRNA expression changes using NanoString nCounter. The heatmap represents the normalized expression of 66 selected circRNAs in ependymomas (blue, n=19), pilocytic astrocytomas (red, n=5), medulloblastomas (purple, n=3) and healthy control samples (green, n=9). Each row in the heatmap corresponds to a unique circRNA and each column corresponds to a patient- or control sample. Each sample is annotated below the dendrogram in the top.

The average circRNA abundance in the ependymoma samples was similar compared to pilocytic astrocytoma and medulloblastoma samples (Fig. 6 and Supplementary Fig. 5). Moreover, all ependymoma samples except one were grouped separately from the pilocytic astrocytoma and medulloblastoma samples with the exception of one pilocytic astrocytoma sample according to principal component analysis (PCA) (Supplementary Fig. 7), indicating that circRNA expression profiles distinguish ependymoma from other pediatric brain tumors.

### Expression levels of individual circRNAs clearly distinguish ependymoma and control samples

In a search for individual circRNAs that may distinguish ependymoma and control samples based on their expression levels, we generated a volcano plot of the NanoString nCounter data and found that many of the differentially expressed circRNAs were highly statistically significant. This mainly applied to the circRNAs downregulated in ependymoma (Fig. 7a). The six significantly upregulated circRNAs in the second cohort of 19 ependymomas and 9 controls were derived from the *PAX3, RMST, LRBA, WDR78, DRC1* and *BBS9* genes, whereas the top 6 most significantly downregulated circRNAs were derived from the *KHDRBS2, ERC2, HOMER1, RIMS1, KCNN2* and *ROCK1* genes (Supplementary Table 11). Among them, circRMST and circRNA derived from *LRBA* gene are differentially expressed in ependymoma only (not in other tumor entities) relative to the control samples (Fig. 7b). Moreover, the circRNAs derived from *ERC2* and *RIMS1* were the only circRNAs downregulated significantly in all the brain tumor samples relative to control samples (Fig. 7b, Supplementary Fig. 8 and Supplementary Table 11).

**Figure 7.**
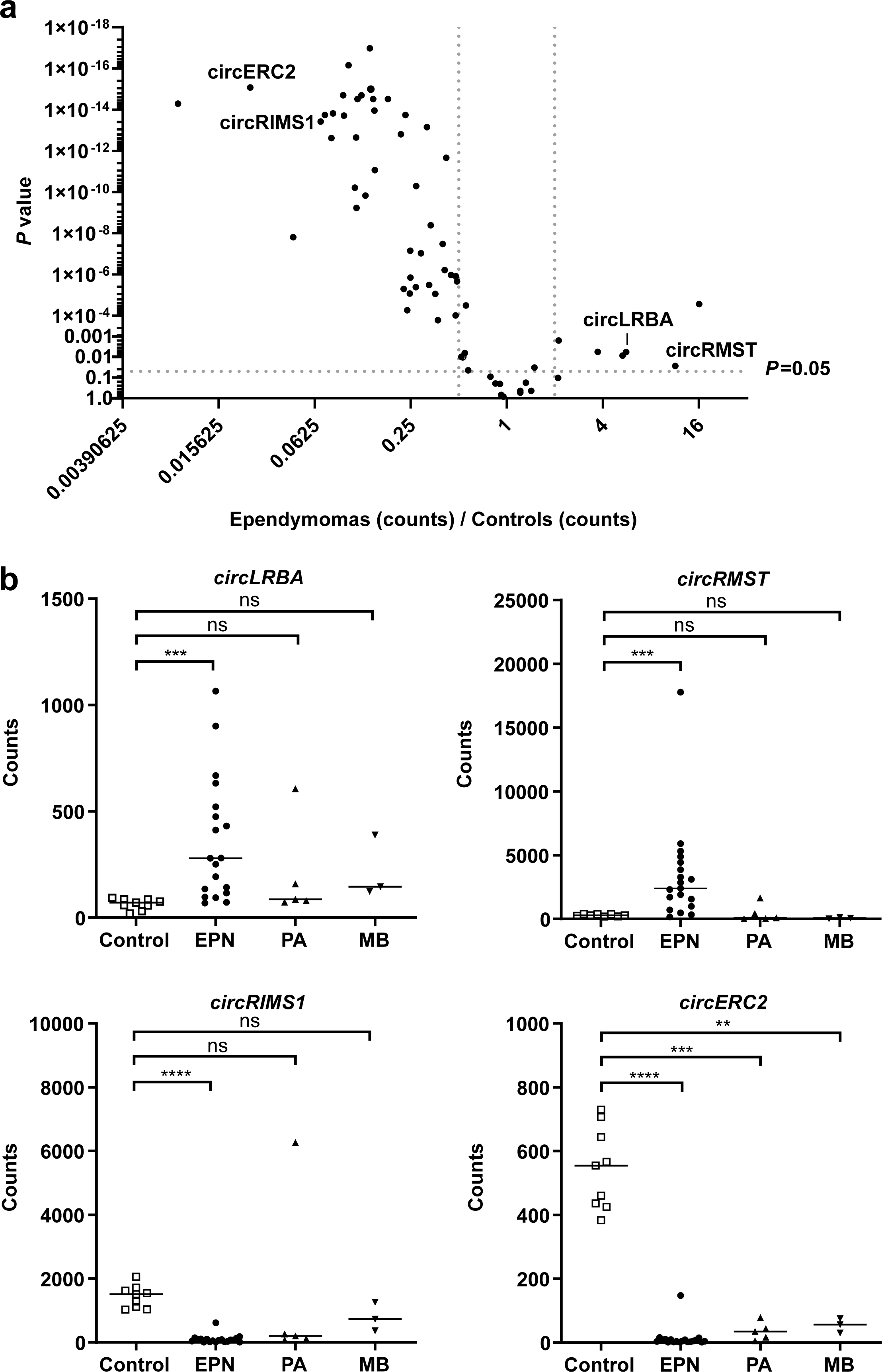
Expression profiling of differentially expressed circRNAs in the NanoString nCounter data. Volcano plot of 66 selected circRNAs showing a high proportion of differentially expressed circRNAs between ependymomas and controls. One unpaired t test for each circRNA was used to generate the p-values. **(b)** Column scatter plots of the expression levels of circRMST and a circRNA derived from *LRBA* gene. EPN: ependymomas (n=19), PA: pilocytic astrocytomas (n=5), MB: medulloblastomas (n=3); Control: healthy control samples (n=9). Mann Whitney U test is used for statistical calculations. * * * *P* < 0.001.

## Discussion

CircRNAs constitute a new class of highly stable endogenous RNAs, mainly thought to have noncoding functions, which are particularly abundant in brain tissues. While numerous studies have already implicated circRNAs as central components in the pathobiology of adult brain tumors, nothing is known about their potential role in pediatric brain tumors. Therefore, we decided to perform high-throughput RNA-sequencing of primary intracranial pediatric ependymomas and control samples. These analyses revealed a high number of differentially expressed circRNAs in the ependymoma samples relative to the control samples, most of which were less abundant in ependymoma. This finding is in line with previous studies indicating that rapidly proliferating cells generally contain fewer circRNAs, possibly due to a dilution effect preventing the highly stable circRNAs from reaching steady-state levels^54-56^.

To our surprise, we also found that circRNA expression profiles derived from unsupervised hierarchical clustering could distinguish ependymoma survivors and patients who died from the disease. We were suprised to see that the overall circRNA expression levels were higher in the deceased patients relative to the survivors as we would expect higher proliferation rates in the more aggressive tumors leading to a greater dilution effect. Thus, we speculate that the deceased patients might have another molecular defect in common, which results in an increased production of circRNAs.

To search for potential trans-acting factors that may drive the observed clustering of ependymoma samples into good and bad prognostic groups, we therefore analyzed a set of genes known to be involved in backsplicing, genes involved in pre-mRNA processing and genes involved in epigenetics. Expression changes in backsplice-promoting (*QKI*^41^, *FUS*^42^ and *ILF3*^43^) and -inhibiting genes (*ADAR*^44^ and *DHX9*^45^) could not explain the observed sub-clustering of the samples. Neither could expression changes in any of 280 genes involved in pre-mRNA processing, although previous research has shown that knockdown of spliceosome genes and splicing factors results in a higher production of circRNAs^46, 57^. Instead, we focused on 167 epigenetic modifier genes and found *DNMT3B, INO80C* and *SETD4* to be significantly more abundant in survivors relative to deceased patients and to correlate with the overall average circRNA expression levels within individual samples in linear regression analyses. *DNMT3B* is responsible for the *de novo* methylation of DNA during early development together with *DNMT3A*^58^, *INO80C* catalyzes ATP-dependent nucleosome sliding and is a component of a chromatin remodeling complex^59^, and *SETD4* is a histone lysine methyltransferase and a member of SET family proteins^60^. Therefore, the combined overexpression of these three genes may have a synergistic effect on circRNA production through epigenetic changes of the host gene bodies, which could potentially explain the observed global differences in circRNA expression. Indeed, the subgroups we identified based on circRNA expression patterns are likely to reflect molecular subgroups defined by 850K methylation analysis, as the PF-EPN-A subgroup is characterized by a lower median age and a dismal outcome^39, 40^. Altogether, we speculate that epigenetic defects in ependymoma not only lead to global changes in DNA methylation, such as CpG island methylator phenotype (CIMP) mainly observed in PF-EPN-A tumors^40^, but also result in global circRNA expression changes. Indeed, the epigenetic states of circRNA host genes (promoters, enhancers and gene bodies) may influence the production of circRNAs^15, 31, 49^. Unfortunately, we were unable to perform 850K DNA methylation analysis to confirm the molecular subgroup of each sample, as the quality of DNA available for these analyses was of too poor. Functional studies of the three genes would be needed to test this hypothesis in the future.

The NanoString nCounter technology is suitable for quantifying circRNAs from FFPE samples based on enzyme-free digital counting^51^. Therefore, we used this technology to validate the circRNA expression patterns of 66 selected circRNAs in ependymoma and to compare with two other pediatric brain tumor entities (pilocytic astrocytoma and medulloblastoma). Most of the circRNA expression changes that were observed using RNA-seq could be reproduced in the NanoString nCounter analysis even though these experiments were performed on another cohort for which only FFPE tissues were available. The heatmap clearly demonstrated that the circRNA expression profile of the ependymoma samples was different from the control samples. Moreover, principal component analysis (PCA) showed that ependymoma samples grouped separately from control samples. This was expected as the circRNAs were selected based on differences between ependymoma- and control samples in the RNA-seq data from the first cohort. In addition, we found that the expression profiles of these circRNAs almost perfectly distinguished the ependymoma samples from pilocytic astrocytoma and medulloblastoma samples despite not being selected for this purpose. Therefore, these preliminary data warrant further investigation into the diagnostic biomarker potential of circRNAs in pediatric brain tumors. In particular, further analysis of individual circRNAs revealed that circRMST and the circRNA derived from the *LRBA* gene are specifically upregulated in the ependymoma samples. In general, we find the upregulated circRNAs to be of particular interest to study further as upregulation, despite the aforementioned dilution effect of circRNAs, may indicate an active selection for these events due to a potential oncogenic function. Nevertheless, many of the downregulated circRNAs are promising as diagnostic or prognostic biomarkers in pediatric brain tumors and should also be investigated further.

## Conclusion

In conclusion, we found a marked global downregulation of circRNAs in ependymoma relative to control samples, and in patients with a good prognosis relative to patients with dismal prognosis. The expression levels of key epigenetic modifier genes correlated with global circRNA abundance in individual samples and we speculate that these genes are responsible for modulating the differences in global circRNA abundance observed between good and poor prognostic groups. In addition, we found that expression levels of individual circRNAs can be used to distinguish ependymoma from healthy brain tissue as well as other pediatric brain tumor entities. Thus, the data presented here suggest that circRNAs could be utilized as diagnostic and prognostic biomarkers in the future if further validated. Finally, future research should aim at investigating the functional relevance of individual deregulated circRNAs in tumorigenesis and progression of pediatric brain cancer.

## Data Availability

Our raw data cannot be submitted to publicly available databases because the patients did not provide consent to share their raw data, which can potentially identify the individuals, but are available from the corresponding author on reasonable request.

## Acknowledgements

We thank Linea Cecilie Melchior for her technical help and Morten Trillingsgaard Venø for his support in bioinformatic analyses. This project was kindly supported by Børnecancerfonden (2016-0176) and Dagmar Marshall Foundation. All authors declare no competing financial interests.

## Supplementary Figure legends

**Supplementary Figure 1. Identification of unique circRNAs supported by at least two sequencing reads (BSJ >2) in ependymoma and control samples. (a-c)** Scatter plots of the top 100 circRNAs (**a**), the top 5000 circRNAs (**b**) and all unique circRNAs (**c**) in ependymoma (n=10) and healthy control (n=3) samples. BSJ: backsplicing junction; RPM: reads-per-million. Mann Whitney U test was used for statistical calculations. * * * * *P* < 0.0001

**Supplementary Figure 2. Expression profiling of backsplicing promoting and inhibiting genes in ependymoma samples. (a)** Column scatter plot of expression profiling of backsplicing promoting genes (*QKI, FUS* and *DHX9*) and inhibiting genes (*ADAR* and *ILF3*) from deceased (red dots, n=5) patients, survivors (blue dots, n=4), and healthy control (green dots, n=3) samples. Mann Whitney U test was used for statistical calculations. **(b)** Correlation between expression of candidate backsplicing modifier genes and circRNA expression (RPM) with corresponding linear regression statistics and R-squared values. Blue dots: samples from survivors; red dots: samples from deceased patients; green dots: samples from healthy controls; purple dot: sample from a patient with an unknown survival status. * * *P* < 0.01, * *P* < 0.05.

**Supplementary Figure 3. Expression profiling of pre-mRNA splicing genes in ependymoma samples**. Scatter plot of the genes associated with pre-mRNA splicing events from deceased (n=5) patients, survivors (n=4), and healthy control (n=3) samples. Mann Whitney U test is used for statistical calculations.

**Supplementary Figure 4. Expression profiling of deregulated pre-mRNA splicing genes in ependymoma samples. (a)** Volcano plot of deregulated genes involved in pre-mRNA splicing events between deceased (n=5) patients and survivors (n=4). One unpaired t test is used to generate the plot. **(b)** Correlation between expression of candidate pre-mRNA splicing genes and circRNA expression (RPM) with corresponding linear regression statistics and R-squared values. Blue dots: samples from survivors; red dots: samples from deceased patients; green dots: samples from healthy controls; purple dot: sample from a patient with an unknown survival status. * * *P* < 0.01, * *P* < 0.05.

**Supplementary Figure 5. Average circRNA expressions of brain tumor entities and healthy control samples from the NanoString nCounter data**. Column scatter plot of average circRNA expression in ependymomas (n=19), pilocytic astrocytomas (n=5), medulloblastomas (n=3) and healthy control samples (n=9). Mann Whitney U test was used for statistical calculations. * * * * *P* < 0.0001.

**Supplementary Figure 6. The correlation between results from the RNA-seq and the NanoString nCounter analyses**. Correlation between log_2_ fold change values between ependymoma samples and control samples in two independent cohorts using two different methods. Fc: fold change.

**Supplementary Figure 7. Principal component analysis (PCA) of the samples used in NanoString nCounter analysis**. PCA plot demonstrating the similarity between samples from ependymoma (n=19), pilocytic astrocytomas (n=5), medulloblastomas (n=3) and healthy control samples (n=9). Red dots: samples from healthy controls; green dots: samples from ependymoma patients; blue dots: samples from medulloblastoma patients; purple dots: samples from pilocytic astrocytoma patients.

## Supplementary Table legends

**Supplementary Table 1**. The list and the expression of the circRNAs selected for the NanoString nCounter analysis. The expression of the circRNAs was obtained from the NanoString nCounter data of ependymoma samples (n=19), pilocytic astrocytoma (n=5), medulloblastoma (n=3) and healthy control (n=3) samples.

**Supplementary Table 2**. The list of the unique circRNAs samples, supported by at least two sequencing reads (BSJ >2), are displayed from RNA-seq analyses of ependymoma (n=10) and control (n=3). BSJ: backsplicing junction.

**Supplementary Table 3**. The list of 1167 high abundance circRNAs quantified by RNA-sequencing of ependymoma (n=10) and control (n=3) samples. RPM: read-per-million.

**Supplementary Table 4**. The list and the expression of the backsplicing promoting (*QKI, FUS* and *DHX9*) and inhibiting (*ADAR* and *ILF3*) genes between ependymoma (n=10) and healthy control samples (n=3). Sample number 1, 5, 8, 9 and 10 are from deceased patients; sample number 2, 3, 6 and 7 are from survivors; sample number 11, 12 and 13 are from healthy control patients; sample number 4 is from a patient with an unknown survival status.

**Supplementary Table 5**. The list and the expression of the genes involved in pre-mRNA processing machinery between ependymoma samples (n=10) and healthy control samples (n=3). Sample number 1, 5, 8, 9 and 10 are from deceased patients; sample number 2, 3, 6 and 7 are from deceased patients; sample number 11, 12 and 13 are from healthy control patients; sample number 4 is from a patient with an unknown survival status.

**Supplementary Table 6**. The list and the foldchanges of the genes involved in the pre-mRNA processing machinery between ependymoma samples (n=10) and healthy control samples (n=3). One unpaired t test is used to calculate the p-values. Correction for multiple testing was performed to calculate adjacent p-values using the Holm-Sidak method.

**Supplementary Table 7**. The list and the expression of the genes involved in the epigenetic modification between ependymoma samples (n=10) and healthy control samples (n=3) and are demonstrated for each sample. Sample number 1, 5, 8, 9 and 10 are from deceased patients; sample number 2, 3, 6 and 7 are from deceased patients; sample number 11, 12 and 13 are from healthy control patients; sample number 4 is from a patient with an unknown survival status.

**Supplementary Table 8**. The list and the fold change of the genes involved in the epigenetic modification between ependymoma samples (n=10) and healthy control samples (n=3). One unpaired t test is used to calculate the p-values. Correction for multiple testing was performed to calculate adjacent p-values using the Holm-Sidak method.

**Supplementary Table 9**. The list of the differentially expressed circRNAs between ependymoma samples (n=10) and healthy control samples (n=3). One unpaired t test is used to calculate the p-values. Correction for multiple testing was performed to calculate adjacent p-values using the Holm-Sidak method. RPM: read-per-million; CTL: circular-to-linear.

**Supplementary Table 10**. The list of the differentially expressed circRNAs between deceased patients (n=5) and survivors (n=4). One unpaired t test is used to calculate the p-values. Correction for multiple testing was performed to calculate adjacent p-values using the Holm-Sidak method. RPM: read-per-million; CTL: circular-to-linear.

**Supplementary Table 11**. Expression levels of 66 circRNAs in ependymoma samples (n=19), pilocytic astrocytoma (n=5), medulloblastoma (n=3) and healthy control (n=3) samples analyzed by the NanoString nCounter. One unpaired t test is used to calculate the p-values. Correction for multiple testing was performed to calculate adjacent p-values using the Holm-Sidak method. RPM: read-per-million; CTL: circular-to-linear; FC: fold change; Epn: ependymoma; PA: pilocytic astrocytoma; MB: medulloblastoma; Control: samples from healthy patients.

## References

1. Gerstner, E.R. & Pajtler, K.W. Ependymoma. Seminars in neurology 38, 104–111 (2018).

2. Pajtler, K.W. et al. The current consensus on the clinical management of intracranial ependymoma and its distinct molecular variants. Acta neuropathologica 133, 5–12 (2017).

3. Ellison, D.W. et al. Histopathological grading of pediatric ependymoma: reproducibility and clinical relevance in European trial cohorts. Journal of negative results in biomedicine 10, 7 (2011).

4. Khatua, S., Ramaswamy, V. & Bouffet, E. Current therapy and the evolving molecular landscape of paediatric ependymoma. European journal of cancer (Oxford, England : 1990) 70, 34–41 (2017).

5. Parker, M. et al. C11orf95-RELA fusions drive oncogenic NF-kappaB signalling in ependymoma. Nature 506, 451–455 (2014).

6. Louis, D.N. et al. The 2016 World Health Organization Classification of Tumors of the Central Nervous System: a summary. Acta Neuropathol 131, 803–820 (2016).

7. Memczak, S. et al. Circular RNAs are a large class of animal RNAs with regulatory potency. Nature 495, 333–338 (2013).

8. Stagsted, L.V., Nielsen, K.M., Daugaard, I. & Hansen, T.B. Noncoding AUG circRNAs constitute an abundant and conserved subclass of circles. Life science alliance 2 (2019).

9. Kristensen, L.S. et al. The biogenesis, biology and characterization of circular RNAs. Nat Rev Genet 20, 675–691 (2019).

10. Kristensen, L.S., Hansen, T.B., Veno, M.T. & Kjems, J. Circular RNAs in cancer: opportunities and challenges in the field. Oncogene 37, 555–565 (2018).

11. Rybak-Wolf, A. et al. Circular RNAs in the Mammalian Brain Are Highly Abundant, Conserved, and Dynamically Expressed. Mol Cell 58, 870–885 (2015).

12. Veno, M.T. et al. Spatio-temporal regulation of circular RNA expression during porcine embryonic brain development. Genome Biol 16, 245 (2015).

13. You, X. et al. Neural circular RNAs are derived from synaptic genes and regulated by development and plasticity. Nat Neurosci 18, 603–610 (2015).

14. Salzman, J., Chen, R.E., Olsen, M.N., Wang, P.L. & Brown, P.O. Cell-type specific features of circular RNA expression. PLoS Genet 9, e1003777 (2013).

15. Enuka, Y. et al. Circular RNAs are long-lived and display only minimal early alterations in response to a growth factor. Nucleic Acids Res 44, 1370–1383 (2016).

16. Jeck, W.R. et al. Circular RNAs are abundant, conserved, and associated with ALU repeats. Rna 19, 141–157 (2013).

17. Hansen, T.B. et al. Natural RNA circles function as efficient microRNA sponges. Nature 495, 384–388 (2013).

18. Abdelmohsen, K. et al. Identification of HuR target circular RNAs uncovers suppression of PABPN1 translation by CircPABPN1. RNA Biol 14, 361–369 (2017).

19. Ashwal-Fluss, R. et al. circRNA biogenesis competes with pre-mRNA splicing. Mol Cell 56, 55–66 (2014).

20. Liu, C.X. et al. Structure and Degradation of Circular RNAs Regulate PKR Activation in Innate Immunity. Cell 177, 865-880.e821 (2019).

21. Hu, D. & Zhang, Y. Circular RNA HIPK3 promotes glioma progression by binding to miR-124-3p. Gene 690, 81–89 (2019).

22. Jin, P. et al. CircRNA circHIPK3 serves as a prognostic marker to promote glioma progression by regulating miR-654/IGF2BP3 signaling. Biochemical and biophysical research communications 503, 1570–1574 (2018).

23. Barbagallo, D. et al. Dysregulated miR-671-5p / CDR1-AS / CDR1 / VSNL1 axis is involved in glioblastoma multiforme. Oncotarget 7, 4746–4759 (2016).

24. Li, X. & Diao, H. Circular RNA circ_0001946 acts as a competing endogenous RNA to inhibit glioblastoma progression by modulating miR-671-5p and CDR1. Journal of cellular physiology 234, 13807–13819 (2019).

25. Zheng, S.Q. et al. CircPCMTD1 Acts as the Sponge of miR-224-5p to Promote Glioma Progression. Frontiers in oncology 9, 398 (2019).

26. Barbagallo, D. et al. CircSMARCA5 Inhibits Migration of Glioblastoma Multiforme Cells by Regulating a Molecular Axis Involving Splicing Factors SRSF1/SRSF3/PTB. Int J Mol Sci 19 (2018).

27. Zhang, M. et al. A peptide encoded by circular form of LINC-PINT suppresses oncogenic transcriptional elongation in glioblastoma. Nature communications 9, 4475 (2018).

28. Zhang, M. et al. A novel protein encoded by the circular form of the SHPRH gene suppresses glioma tumorigenesis. Oncogene 37, 1805–1814 (2018).

29. Yang, Y. et al. Novel Role of FBXW7 Circular RNA in Repressing Glioma Tumorigenesis. J Natl Cancer Inst 110 (2018).

30. Zhang, X.O. et al. Complementary sequence-mediated exon circularization. Cell 159, 134–147 (2014).

31. Kristensen, L.S., Okholm, T.L.H., Veno, M.T. & Kjems, J. Circular RNAs are abundantly expressed and upregulated during human epidermal stem cell differentiation. RNA Biol 15, 280–291 (2018).

32. Liao, Y., Smyth, G.K. & Shi, W. featureCounts: an efficient general purpose program for assigning sequence reads to genomic features. Bioinformatics 30, 923–930 (2014).

33. Tang, C. et al. Template switching causes artificial junction formation and false identification of circular RNAs. bioRxiv, 259556 (2018).

34. Szabo, L. & Salzman, J. Detecting circular RNAs: bioinformatic and experimental challenges. Nat Rev Genet 17, 679–692 (2016).

35. Glazar, P., Papavasileiou, P. & Rajewsky, N. circBase: a database for circular RNAs. RNA 20, 1666–1670 (2014).

36. Dong, R., Ma, X.K., Li, G.W. & Yang, L. CIRCpedia v2: An Updated Database for Comprehensive Circular RNA Annotation and Expression Comparison. Genomics Proteomics Bioinformatics 16, 226–233 (2018).

37. Yang, P. et al. Silencing of cZNF292 circular RNA suppresses human glioma tube formation via the Wnt/beta-catenin signaling pathway. Oncotarget (2016).

38. Song, X. et al. Circular RNA profile in gliomas revealed by identification tool UROBORUS. Nucleic Acids Res 44, e87 (2016).

39. Pajtler, K.W. et al. Molecular Classification of Ependymal Tumors across All CNS Compartments, Histopathological Grades, and Age Groups. Cancer Cell 27, 728–743 (2015).

40. Mack, S.C. et al. Epigenomic alterations define lethal CIMP-positive ependymomas of infancy. Nature 506, 445–450 (2014).

41. Conn, S.J. et al. The RNA binding protein quaking regulates formation of circRNAs. Cell 160, 1125–1134 (2015).

42. Errichelli, L. et al. FUS affects circular RNA expression in murine embryonic stem cell-derived motor neurons. Nat Commun 8, 14741 (2017).

43. Li, X. et al. Coordinated circRNA Biogenesis and Function with NF90/NF110 in Viral Infection. Mol Cell 67, 214–227 e217 (2017).

44. Ivanov, A. et al. Analysis of intron sequences reveals hallmarks of circular RNA biogenesis in animals. Cell Rep 10, 170–177 (2015).

45. Aktas, T. et al. DHX9 suppresses RNA processing defects originating from the Alu invasion of the human genome. Nature 544, 115–119 (2017).

46. Liang, D. et al. The Output of Protein-Coding Genes Shifts to Circular RNAs When the Pre-mRNA Processing Machinery Is Limiting. Mol Cell 68, 940-954.e943 (2017).

47. Sveen, A. et al. Transcriptome instability in colorectal cancer identified by exon microarray analyses: Associations with splicing factor expression levels and patient survival. Genome medicine 3, 32 (2011).

48. Singh Nanda, J., Kumar, R. & Raghava, G.P. dbEM: A database of epigenetic modifiers curated from cancerous and normal genomes. Sci Rep 6, 19340 (2016).

49. Ferreira, H.J. et al. Circular RNA CpG island hypermethylation-associated silencing in human cancer. Oncotarget 9, 29208–29219 (2018).

50. Fagerberg, L. et al. Analysis of the human tissue-specific expression by genome-wide integration of transcriptomics and antibody-based proteomics. Molecular & cellular proteomics : MCP 13, 397–406 (2014).

51. Dahl, M. et al. Enzyme-free digital counting of endogenous circular RNA molecules in B-cell malignancies. Lab Invest 98, 1657–1669 (2018).

52. Chen, D.-F., Zhang, L.-J., Tan, K. & Jing, Q. Application of droplet digital PCR in quantitative detection of the cell-free circulating circRNAs. Biotechnology & Biotechnological Equipment, 1–8 (2017).

53. Conn, V. & Conn, S.J. SplintQuant: a method for accurately quantifying circular RNA transcript abundance without reverse transcription bias. Rna 25, 1202–1210 (2019).

54. Bachmayr-Heyda, A. et al. Correlation of circular RNA abundance with proliferation--exemplified with colorectal and ovarian cancer, idiopathic lung fibrosis, and normal human tissues. Sci Rep 5, 8057 (2015).

55. Moldovan, L.I. et al. High-throughput RNA sequencing from paired lesional- and non-lesional skin reveals major alterations in the psoriasis circRNAome. BMC Med Genomics 12, 174 (2019).

56. Das Mahapatra, K. et al. A comprehensive analysis of coding and non-coding transcriptomic changes in cutaneous squamous cell carcinoma. Sci Rep 10, 3637 (2020).

57. Kramer, M.C. et al. Combinatorial control of Drosophila circular RNA expression by intronic repeats, hnRNPs, and SR proteins. Genes Dev 29, 2168–2182 (2015).

58. Gagliardi, M., Strazzullo, M. & Matarazzo, M.R. DNMT3B Functions: Novel Insights From Human Disease. Front Cell Dev Biol 6, 140 (2018).

59. Jin, J. et al. A mammalian chromatin remodeling complex with similarities to the yeast INO80 complex. J Biol Chem 280, 41207–41212 (2005).

60. Faria, J.A. et al. SET domain-containing Protein 4 (SETD4) is a Newly Identified Cytosolic and Nuclear Lysine Methyltransferase involved in Breast Cancer Cell Proliferation. J Cancer Sci Ther 5, 58–65 (2013).

